# *Spaces*: A Student-Informed, Course-Based Program to Enhance Wellbeing and Present-Moment Awareness in University Undergraduates

**DOI:** 10.64898/2026.01.14.26344125

**Authors:** Donald J. Noble, Charles L. Raison

## Abstract

**Introduction:** The wellbeing of university undergraduates is a pressing area of concern, with one in five students reporting serious psychological distress. In previous semesters of a course on mental wellbeing, we asked undergraduates to engage with a standardized wellness program for the general population and provide feedback through weekly surveys and a final reflection paper. Based on these responses, we developed the 30-day, course-embedded *Spaces* program to more specifically address college student needs.

**Methods:** The *Spaces* program incorporates intrinsically pleasurable activities and short presence practices. We piloted the program in a cohort of 30 students during the Fall 2024 semester. Students completed weekly surveys measuring anxiety, depression, and wellbeing, pre-post surveys on present-moment awareness, and a post-program engagement scale.

**Results:** Over the five program weeks, students in the *Spaces* semester experienced significantly less anxiety (F_(1,4)_=38.40, *P*=0.003) and depression (F_(1,4)_=32.87, *P*=0.005) and more wellbeing (F_(1,4)_=65.86, *P*=0.001) than those in semesters with the standardized program, and increased present-moment awareness compared to baseline (t_(27)_=3.859, *P*=0.0006). Engagement with specific components predicted improvements in mood and attention.

**Discussion:** These findings highlight the potential of an iterative approach that incorporates input from the target population in enhancing the ability of a wellness program to meet its goals.

## 1 Introduction

Recent data point to a global reduction in happiness and life satisfaction throughout young adulthood compared to previous decades (VanderWeele et al., 2025). This effect may be especially pronounced in college settings, where 20% of students experience serious psychological distress (ACHA, 2025) and more than one in three report moderate anxiety or depression (HMN, 2025). Unfortunately, wellness interventions have mostly failed to address this problem in college-age students due to multiple factors including lack of flexibility or personalization in the interventions, low faculty involvement, and failure to frame benefits in a comprehensible scientific context (Dajani et al., 2021). For example, a cluster randomized controlled trial of more than 8,000 adolescents across 84 schools found no improvements in depression or wellbeing with school-based mindfulness training compared to standard social-emotional teaching (Kuyken et al., 2022). The authors concluded that “There is need to ask what works, for whom and how, as well as considering key contextual and implementation factors.” Indeed, there is increasing recognition of the need to identify specific components (“active ingredients”) of mental health interventions that are effective for alleviating mental health symptoms in young people as an important initial step on the road to developing more targeted and effective treatments.

Over five years we have offered an empirically-supported, manualized wellness program as an experiential component in a college course on wellbeing. This program focuses on enhancing five pillars of wellness, including mindfulness, sleep, exercise, nutrition and social connectedness (Rolin et al., 2020; Jain, 2023). While students frequently noted benefits from participating in the program, they also found it challenging to incorporate all five elements consistently into their daily lives. As a result, students often chose just one or two of the five modules to focus on. Qualitatively (and quantified as part of the present study), we did not observe improvements in scores on the weekly anonymous anxiety, depression, and wellbeing scales completed during the course. In each of the semesters this program was implemented, students also submitted final reflection papers identifying their perceptions of the strengths and limitations of the standardized program. These reflections inspired us to begin an iterative process with the students to identify program elements that might be modified to address commonly cited limitations impeding uptake of the wellness program. One pervasive pattern that emerged was an inability/unwillingness on the part of students to engage with the standardized instructions of the program. We also asked students to suggest other elements they would like to see in a wellness program. We decided to take a lesson from both these streams of feedback and design a new program that incorporated the most commonly requested elements while remaining flexible enough to be pragmatic for busy college lifestyles.

Over the five semesters that we offered the ready-made wellness program, the five most commonly suggested elements students wanted to see added, which form the core of our *Spaces* program, are Nature, Spirituality & Resonance Breathing, Adaptive Stress, Creative Hobbies, and Quality Relationships. Scientific data support the potential of each element to enhance wellbeing (Bernardi et al., 2001; Stuckey and Nobel, 2010; Bratman et al., 2015; Janssen et al., 2016; Noble and Hochman, 2019; Fincham et al., 2023; Castelo et al., 2025). We incorporated short presence practices (micropractices) into the program as a way for students to engage with the core activities (or, optionally, their everyday routines). For example, students could embed short periods of ‘pure perception’ during a 30-minute walk in nature. This approach sought to better fit the program into students’ existing schedules, amplifying their motivation and willingness to commit time; essentially, making the program more doable. This strategy finds support from leading meditation researchers who increasingly recommend shorter bouts of focused meditation or the incorporation of meditative techniques into everyday life (Brintz et al., 2024; Schwerdtfeger et al., 2025). For example, a randomized controlled trial of mostly public school teachers found that meditating for only 5.5 minutes each day, on average, led to robust reductions in distress within one week compared to a well-matched control group (Hirshberg et al., 2022). The *Spaces* program offered a means to incorporate short presence practices directly into the core program activities, matching the practices to pleasurable sensations to increase student motivation and reinforce daily practices based on principles of behavioral conditioning (Noble et al., 2017). Along with the micropractices, student self-determination was critical for crafting a program workable for each student. We encouraged students to focus on activities they spontaneously enjoyed and that built upon their existing routines or goals; for example, using the program as justification to complete a piece of artwork they had been putting off because it conflicted with academic obligations. Two illustrative reflections documenting student experiences with the program are shown in **Box 1**.

We deployed the program in a pilot cohort of college undergraduate students during the Fall 2024 semester. For comparison, a demographically similar control cohort comprising students from four previous academic semesters and one subsequent semester was analyzed over identical weekly time points in those semesters. We sought to determine i) whether the *Spaces* program cohort experienced significant improvements in mental health over 30 days, and ii) whether engagement with specific program components predicted subsequent changes in anxiety, depression, wellbeing, and present-moment awareness.

### Box 1.

**Example reflections from students in the *Spaces* cohort**

**Student 1:** “When experiencing the walks at first it was somewhat difficult to focus on the environment because as a college student, we tend to have our headphones listening to music wherever we walk… When taking a walk for the first time without listening to music, it made me feel quite anxious when walking around Lullwater, only lasting less than ten minutes then I decided to use my AirPods… Then the next day I tried walking to class without music and while it was a bit frightening, I was able to do so. During my walk to class, I was not able to focus on my surroundings much because I was just trying to rush to class with the awkwardness that I did not have music playing. Then I tried again to walk at Lullwater, but this time I purposely left my AirPods, so I would not be able to use them, and it was somewhat successful. Through this walk, it was challenging at first to not try to daydream or think about things I had to do after, but then I was able to clear my brain to just concentrate on the environment. It was very overwhelming at first, but I was able to take in all the nature that I was seeing. I noticed for the first time that Fall was *here*. I was able to notice the leaves changing colors, the trees, the weird bird standing in the middle of the lake, things that I would not notice when listening to music or when not walking with the intent of paying attention. I continued to do these walks, noticing that I was less anxious about them and more relaxed and more focused on my surroundings, and eventually, through the 30 days, I genuinely felt more at ease with myself.”

**Student 2:** “A particular habit I was able to sustain over the 30-day program was under the quality relationships module; I called my family members and friends twice a week. Although this is something I often did prior to the program, what I changed was I could not treat it as a secondary thing. I used to call people while working on an assignment or cleaning, having the conversations more as background; for these conversations to become intentional, I avoided these distractions. When talking to my great grandmother on the phone, I realized I could hear the background noises which were characteristic of my childhood—the television running on, her runny nose, the creaky ceiling fan. It was powerful enough to transport me back there, closing the gap that thousands of miles of distance had opened. Listening with my whole body changed the way I communicated both over the phone and in-person. I often became teary-eyed when talking to relatives, reminding me of the love I continue to hold for them even from far away. Allowing myself to feel these sensations rather than suppressing them meant recognizing my emotions as they happened rather than in retrospect, enhancing my in-person conversations in which I found myself much more connected to others.

Changes did not have to be extravagant—I have tried that and often lost interest, and even money. Altering an already existing habit that is close to the values that guide me through life was much more effective.”

## 2 Methods

### Study Overview

Data from the current study were initially collected for course quality improvement purposes. Because of this, the study was not pre-registered. Recognizing upon completion of data collection that results were highly relevant to the impact of course content on student wellbeing, study authors petitioned the Emory University Institutional Review Board (IRB) and the study was reviewed and deemed exempt from IRB approval.

### Participants

30 Emory University undergraduate students who enrolled in the course titled “21st Century HLTH & Wellbeing: Ancient Practices Meet Modern Science” in Fall (F) 2024 were included in this longitudinal observational study. Twenty-eight of the students provided complete datasets and were included in within-cohort analyses (see below). For comparison, 270 additional Emory University undergraduates enrolled in five semesters of a complementary course called “Mental Wellbeing & Resilience” over five semesters (F20, F21, Spring [S] 2022, S24, and S25) were analyzed over equivalent time points in the relevant semesters. The breakdown of different class years (Freshmen through Seniors), ethnicities, and legal sexes is reported for these two groups in **Table S1** in the Supplementary Material.

### Wellness Programs

In the F24 semester, students were introduced in class to a program titled *Creating Space for Health* (i.e., *Spaces*) and provided with a workbook. Program design was informed by student input, with instructors (DJN and CLR) choosing the best answers to the prompt “what is missing from your wellness routine outside of the conventional areas of exercise, nutrition, social connectedness, mindfulness meditation, and sleep?” from more than 343 reflection papers submitted during prior semesters. Program modalities (**Table 1**) were chosen based on approximate frequency of suggestion and available scientific support for proposed practices. The program asked students to engage with each of five core activities and two out of five presence micropractices each day for 30 days. During live classes, students were encouraged to experiment with the activities and practices they found most useful and commit to a particular plan for 30 days, rather than rigidly adhere to the full program. Engagement with the program was encouraged but not required, with grading assessment contingent on the quality of a final reflection paper describing experiences and limitations of the program, not on engagement with the program per se. Semester weeks 8 and 13 of the course were the start and endpoint of the program, respectively. Students in the control group (from other semesters) engaged with a preexisting wellness program focused on exercise, mindfulness, nutrition, sleep, and social connectedness (Rolin et al., 2020; Jain, 2023).

**Table 1.** Summary of *Spaces* Program Components.

|  |
| --- |
| **Module 1. Nature**<br>**Activity:** Spend 20 minutes outdoors appreciating the sensations of nature.<br>**Ideas:** • Go for a nature walk • Stargaze at night • Spend time outside with an animal • Take a detour and go the long way between classes • Find a “thin place” where you can practice pure perception every day<br>**Presence Micropractice:** Exteroception (Observing Outer Sensations)<br>2-3 times per day, spend at least 20 seconds in a state of pure sensory perception. |
| **Module 2. Spirituality & Resonance Breathing**<br>**Activity:** Spend 10 minutes combining a chosen spiritual practice with resonance breathing.<br>**Ideas:** • Read inspiring nonfiction while breathing deeply • Pray during deep inhales • Recite the Ave Maria or a yogic mantra (“automatic” slow breathing) • Meditate to the recording of the chimes or a sound bath<br>**Presence Micropractice:** Interoception (Observing Inner Sensations)<br>Each day, take 3 mindful breaths or spend 30s – 1min scanning your body. |
| **Module 3. Adaptive Stress**<br>**Activity:** Try out one deprivation-based adaptive stressor.<br>**Ideas:** • Try Wim Hof high-ventilation breathwork • Choose between or alternate hot baths and cold showers • Exercise using high-intensity interval training • Cut out sugar, carbs, and alcohol, or attempt mindful fasting<br>**Presence Micropractice:** Meta-awareness (Observing Your Mind)<br>Each day, pause 3 times to observe and record your thoughts and emotions. |
| **Module 4. Creative Hobbies**<br>**Activity:** Spend 15 minutes completely immersed in an activity.<br>**Ideas:** • Jog, play tennis, or stretch • Engage with dance, yoga, or spontaneous movement practices • Write poetry, draw, paint, or try theatre • Try out horticulture or community gardening<br>**Presence Micropractice:** Mindful Action (Attention to Movement)<br>Each day, give your complete attention to a routine task. |
| **Module 5. Quality Relationships**<br>**Activity:** Limit your social media usage and engage in one intentional social interaction.<br>**Ideas:** • Only access social media in the morning and evening • Do not access social media in the presence of others • Have a conversation with the dining hall staff every day • Give your attention fully to someone in the elevator<br>**Presence Micropractice:** Mindful Listening (Attention to Others)<br>Each day, spend at least 20 seconds giving someone your full attention. |

### Anxiety, Depression, Wellbeing, Presence, and Engagement Surveys

Students were notified in class by course instructors prior to the end of the enrollment period that their enrollment in the course constituted an agreement to anonymously fill out the weekly surveys. Students anonymously completed clinically validated scales measuring anxiety (7-item Generalized Anxiety Disorder Scale [GAD-7]) (Spitzer et al., 2006), depression (2-Item Patient Health Questionnaire [PHQ-2]) (Kroenke et al., 2003), and general wellbeing (World Health Organization-Five Wellbeing Index [WHO5]) (Topp et al., 2015) each week throughout the 13-14 week course, as well as a custom-made 7-item Presence scale – with several questions adapted from a recently validated research inventory (Kilrea et al., 2023) – on Weeks 8 (before program) and 13 (program endpoint), and a survey measuring program engagement on Week 13. No engagement data were collected for the control semesters. Survey questions, answer options, and recommended cutoffs for further mental health screening are shown in **Table S2** in the Supplementary Material. Total possible scores on the Presence survey ranged from 7 to 35 (higher = more presence), on the GAD-7 ranged from 0 to 21 (higher = more anxious), on the PHQ-2 from 0 to 6 (higher = more depressed), and on the WHO5 from 0 to 25 (higher = greater wellbeing). Responses were submitted online through the Canvas Learning Management System, via Likert-type scales. Surveys were typically open for 5-7 days each week and closed on Wednesday night (most semesters) or Thursday morning (F20). While we did not directly screen students for DSM-5 depressive or anxiety disorders, each of the scales provides recommendations on the thresholds for further clinical screening. Links to university and external mental health resources and contact information were provided on Canvas and mentioned once per week in class. School credit (1 class participation point) was awarded for completion of each survey.

### Data Analysis

Analyses were performed separately for each scale since they are designed to measure different aspects of overall wellbeing and because they have different maximal scores. Weekly survey averages (total scores) were organized by group. These values were averaged across weeks to derive overall mean and standard deviation values for each group (**Table S1** in the Supplementary Material). To evaluate group differences in mental health over time while controlling for baseline levels, we fit a weighted least-squares (WLS) general linear model with Group (Spaces versus Control) and Week (1-5) as main effects for each outcome scale. The dependent variable was score change from baseline at each program week (Δ = post – baseline). To account for differences in sample size and variability across timepoints, each value was weighted by the inverse variance of Δ, so weeks with larger N and smaller SD contributed more. The model yielded a week-adjusted Spaces – Control difference in change. Because individuals were anonymous (no individual-level pairing), baseline and weekly samples were treated as independent when computing the variance of Δ; this likely overestimates variance (i.e. conservative p-values). As an exploratory test, we repeated the model treating Week as numeric and added a Group×Week interaction (1-df linear trend) to test whether weekly slopes of change differed between groups. Subsequent analyses were restricted to the F24 *Spaces* program semester. For this cohort, data were collected anonymously but Canvas maintained consistent identifiers for “Student 1”, “Student 2”, etc., permitting repeated-measures analysis on a within-subjects basis. For the comparison cohorts (F20, F21, S22, S24, and S25), only aggregate data were available. For the Presence scale, on which only baseline and post-program responses were obtained, a paired t-test was run to determine changes in present-moment awareness before vs. after the program. To understand potential predictors of outcomes (changes in mental health), correlation analyses with subsequent linear regression were performed for each scale with levels of engagement (with each program component and overall) and changes in wellness scores from Baseline to the endpoint of the program (five weeks) preselected as variables of interest. Correlations were performed using weekly data, rather than semesterly averages. To avoid statistical over-testing (i.e. multiple comparisons problem), we performed targeted comparisons between program components and overall survey scores (GAD-7, PHQ-2, WHO5), only performing sub-question analysis if results were significant or at the threshold of significance for scale totals. Sub-questions from the Presence scale were individually analyzed to determine whether engagement with individual micropractices predicted improvements in different modalities of presence (e.g. focused attention versus peace and harmony). Statistical analyses were performed using IBM SPSS Statistics version 31 and GraphPad Prism version 10.6 software. Data are presented as mean ± SD unless otherwise indicated, with two-tailed tests and significance set at *P* ≤ .05.

## 3 Results

### 3.1. Changes in mental health during the program period

Scores on each of the three wellness scales (GAD-7, PHQ-2, and WHO5) were analyzed over the program weeks as changes from baseline (Δ = post – baseline), using weighted least-squares (WLS) general linear models to assess main effects of Group and Week **(Figure 1)**.

**Figure 1.**
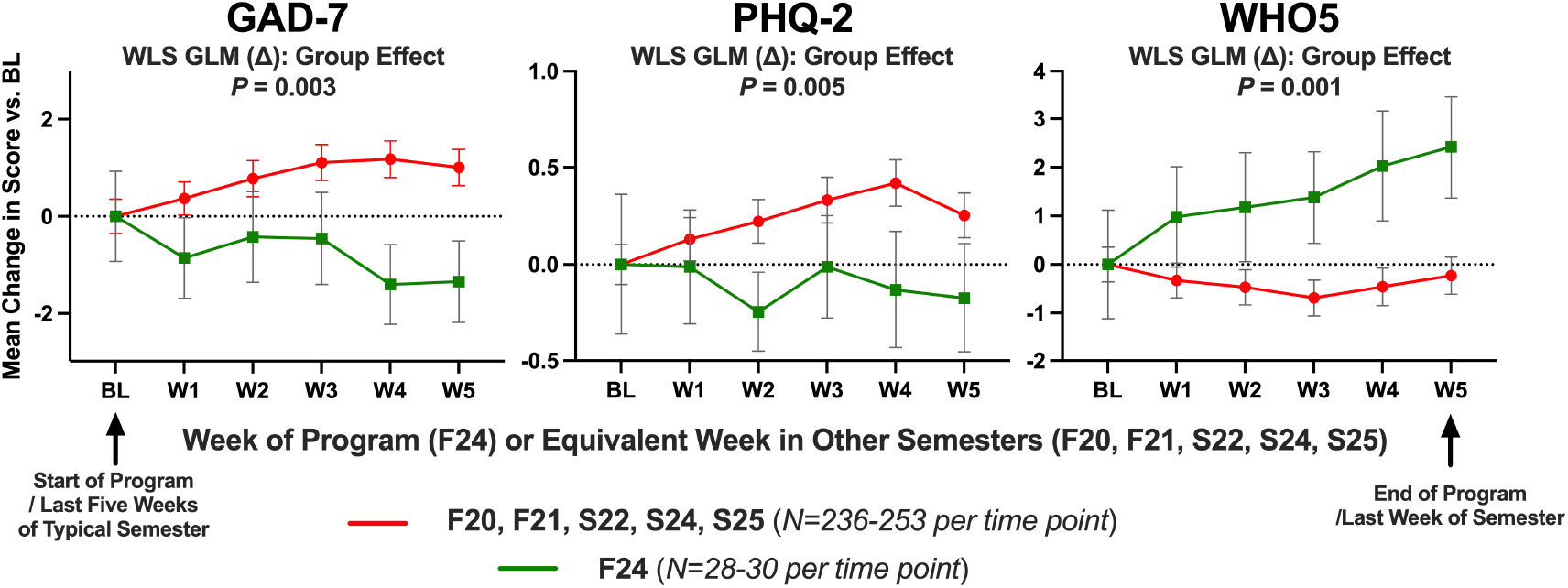
Improved Mental Health Over 30-Day *Spaces* Program. Lines show weekly changes from baseline (Δ) as mean ± SEM for the *Spaces* (*green*) and control (*red*) cohorts. Statistics are from weighted least-squares general linear models on Δ with Group and Week as main effects. Students in the *Spaces* program cohort experienced improved anxiety (GAD-7), depression (PHQ-2), and wellbeing (WHO5) over the five weeks of the program compared to a control cohort of students from five other semesters who were exposed to a more standardized wellness program. The *Spaces* cohort also showed a linear increase in wellbeing over time compared to the control cohort (*P* = 0.05).

#### GAD-7

The adjusted mean change was −0.91 (95% CI −1.66 to −0.17) in the *Spaces* program group and 0.88 (95% CI 0.59 to 1.18) in the control group. The model explained a substantial proportion of variance (*R*^*2*^ = .917, adj. .814). The *Spaces* vs. control cohort difference was significant (F_(1,4)_= 38.40, *P* = 0.003; *Spaces* – control adjusted difference −1.80 points, 95% CI −2.60 to −1.00). A Group×Week linear trend suggested borderline faster improvement for the *Spaces* group (≈-0.36 points/week; *P* = 0.067).

#### PHQ-2

The adjusted mean change was −0.12 (95% CI −0.30 to 0.06) in the *Spaces* program group and 0.27 (95% CI 0.21 to 0.33) in the control group. The model accounted for a substantial proportion of variance in change scores (*R*^*2*^ = .927, adj. .836). The *Spaces* vs. control cohort difference was significant (F_(1,4)_= 32.87, *P* = 0.005; *Spaces* – control adjusted difference −0.39 points, 95% CI −0.58 to −0.20). There was no Group×Week linear trend (*P* = 0.434).

#### WHO-5

The adjusted mean change was 1.60 (95% CI 0.94 to 2.25) in the *Spaces* program group and −0.44 (95% CI −0.66 to −0.21) in the control group. The model explained a large proportion of variance (*R*^*2*^ = .946, adj .880). The *Spaces* vs. control cohort difference was significant (F_(1,4)_= 65.86, *P* = 0.001; *Spaces* – control adjusted difference 2.03 points, 95% CI 1.34 to 2.72). The Group×Week linear trend indicated faster gains for the *Spaces* group (≈0.35 points/week; *P* = 0.050).

There was no effect of Week for any of the outcomes measured.

### 3.2. Associations between program engagement and mental health outcomes

Pearson correlation coefficients were determined between self-reported engagement with the program and mental health outcomes. Refer to **Table 1** for further details on the core program modules and presence micropractices. Survey instruments are reported in **Table S2** in the Supplementary Material.

Total time engaging with the program did not correlate with mental health outcomes (GAD-7: *P* = 0.81; PHQ-2: *P* = 0.58; WHO5: *P* = 0.42). Instead, engagement with specific activities and practices predicted changes in mental health.

Correlations between program activities and mental health:

#### Baseline

Students who went on to engage more with *Nature* tended to be less anxious overall before beginning the program (*r* = −0.37, *P* = 0.053). Sub-question analysis revealed that they rated themselves as having less trouble relaxing (*r* = −0.50, *P* = 0.0063) at baseline. Students who chose to engage more with *Intentional Interactions* also tended to be less anxious overall prior to the program (*r* = −0.37, *P* = 0.052), being less restless (*r* = −0.44, *P* = 0.021) and less afraid of something awful happening (*r* = − 0.39, *P* = 0.042). Baseline depression and wellbeing scores did not predict subsequent engagement with program activities.

#### Pre-post

Students who engaged more with *Spirituality and Resonance Breathing* saw an overall improvement in wellbeing after the program (*r* = 0.38, *P* = 0.044; **Figure 2A**). Sub-question analysis revealed they had less trouble relaxing (*r* = −0.40, *P* = 0.035) and were more calm/relaxed (*r* = 0.39, *P* = 0.041) and fresh/rested (*r* = 0.48, *P* = 0.0099). Paradoxically, students who engaged more with *Intentional Interactions* felt more depressed overall (*r* = 0.41, *P* = 0.030; **Figure 2B**), with sub-questions indicating these students felt more down/hopeless (*r* = 0.42, *P* = 0.025). There were no pre-post associations with anxiety.

**Figure 2.**
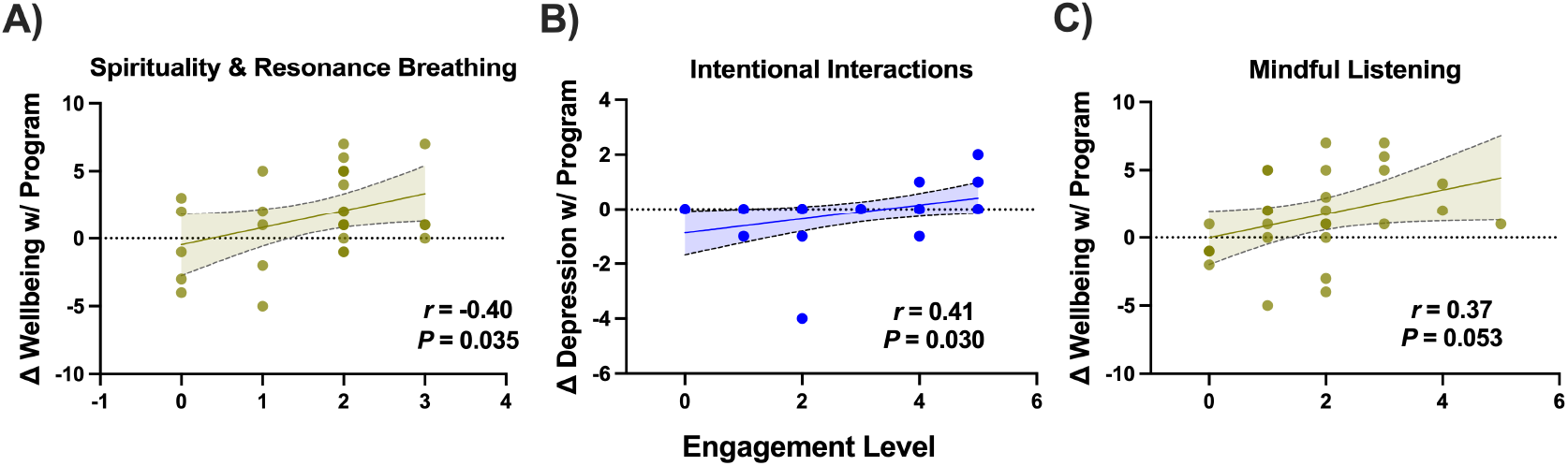
*Spaces* Program Engagement Predictors of Mental Health Outcomes. (**A**) The program module on spirituality and resonance breathing predicted overall improvements in wellbeing within the *Spaces* cohort. (**B**) Unexpectedly, students who engaged the most with a modular activity focused on the *quantity* of social interactions (adding one interaction each day) felt more depressed after the program. (**C**) Focusing more on the *quality* of social interactions through mindful listening (a presence micropractice) predicted positive changes in wellbeing at the threshold of statistical significance. *In A-C, Δ wellbeing or Δ depression indicate pre-post changes in WHO5 or PHQ-2 scale totals, respectively; darker coloured circles indicate multiple students for a given engagement-Δ mood combination*.

Correlations between presence micropractices and mental health:

#### Baseline

Students who chose to engage more frequently with *Interoception* reported feeling more overall depression (*r* = 0.38, *P* = 0.044), including less interest/pleasure in doing things (*r* = 0.43, *P* = 0.021), before beginning the program. Baseline anxiety and wellbeing scores did not predict subsequent engagement with any of the presence micropractices.

#### Pre-post

Individuals who engaged more with *Mindful Listening* tended to experience increased overall wellbeing after finishing the program, with a positive correlation on the threshold of statistical significance (*r* = 0.37, *P* = 0.053; **Figure 2C**). These students reported that their lives were more filled with interesting things (*r* = 0.49, *P* = 0.0084). There were no pre-post associations with anxiety or depression.

### 3.3. Changes in presence and associations with program engagement

We also analyzed results from a custom scale intended to measure present-moment awareness, which students completed before and immediately after the program (Weeks 8 and 13 on the semester timeline).

Student total scores on the 7-item Presence survey significantly increased from baseline to post-program (paired t-test, t_(27)_= 3.859, *P* = 0.0006; **Figure 3A**). Sub-questions were analyzed to isolate effects on different modalities of presence. As shown in **Figure 3B**, students who reported the greatest increases in sustained attention were those who engaged most with *Exteroception* (*r* = 0.58, *P* = 0.001) and *Interoception* (*r* = 0.59, *P* = 0.001). Students who engaged more with *Interoception* also reported feeling an increased sense of peace after completing the program (*r* = 0.43, *P* = 0.024).

**Figure 3.**
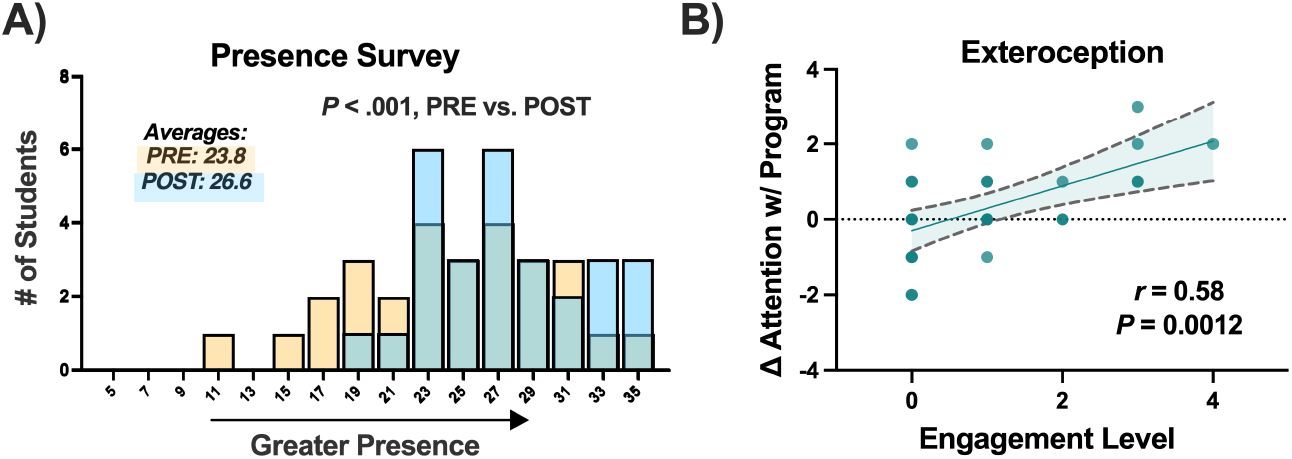
Presence Scale and Engagement Predictors. A custom scale (Presence) was constructed to measure pre-post changes in present-moment awareness. **(A)** Frequency histograms showing individual student scores before and after the *Spaces* program, with semi-transparent bars (i.e. green indicates areas of overlap between pre [*yellow*] and post [*blue*] bars). Total scores on this scale were significantly higher after completing the program. **(B)** Correlations between micropractices and present-moment awareness revealed that engagement with a micropractice focused on exteroception (awareness of outer sensations) strongly predicted a positive change in students’ ability to focus attention for long periods of undistracted time after completing the program. A similar result was observed for interoception (awareness of inner sensations).

An exploratory analysis of engagement profiles corresponding to mental health outcomes is presented in **Result S1** and **Figure S1** in the Supplementary Material.

## 4 Discussion

We deployed a student-informed wellness program, *Spaces*, in a pilot cohort of 30 college undergraduates. Student anxiety and depression levels decreased and wellbeing increased over the 30-day program compared to a demographically similar control cohort that attempted a program focused on basic wellness behaviors for the general population. The *Spaces* cohort also reported an overall increase in present-moment awareness immediately after completing the program. Engagement with specific elements, rather than the program as a whole, differentially predicted mental health outcomes. Findings from our pilot study support implementation of personalized, multi-component wellness interventions that respond directly to the needs of a target population.

The *Spaces* program had a positive effect on subjective mental health compared to the control condition, as captured by the GAD-7, PHQ-2, and WHO5 scales. The WHO5 effect was particularly striking, with an adjusted mean change of 1.60 points and linear trend supporting a 0.35 points/week improvement in the *Spaces* cohort. Based on these results, we sought to address whether engagement with specific components of the program predicted changes in mental health. Correlation analysis with subsequent linear regression revealed that greater student engagement with the program’s spirituality and resonance breathing activity predicted improvements in wellbeing. Unexpectedly, students who engaged in intentional interactions – characterized by overt attempts to engage with at least one individual per day – felt more depressed by the end of the program. This finding seemingly contradicts the well-documented ability of strong and supportive social relationships to increase an individual’s happiness (Waldinger and Schulz, 2023). However, we also found that students who engaged more with mindful listening – committing to at least 20 seconds per day of giving someone their full attention – tended to experience greater wellbeing after the program. One of our students noted that “my emotional satisfaction doesn’t necessarily increase with more frequent social encounters. In fact, I often derive greater value from fewer, but more substantive, gatherings… promoting genuine wellbeing through sustainable and meaningful practices.” Thus, it may be that bringing quality to existing social interactions is more important than increasing their quantity, a finding that if replicated in larger prospective studies might open new perspectives on how to optimize social engagement strategies in young people.

Interestingly, total time spent on the program did not associate with improved mental health. Rather, more frequent engagement with shorter program components associated with pre-post improvements in wellbeing. For example, the spirituality and resonance breathing activity encouraged students to enhance the quality of preexisting religious rituals like prayer or secular activities such as nonfiction reading. In addition to spirituality and resonance breathing, this included a short presence practice focused on mindful listening. On average, students spent less time engaging with these elements than most other parts of the program, yet the students who did engage more tended to feel better. This finding extended to a 7-item scale that we designed and administered at pre-post timepoints to assess present-moment awareness. The program cohort reported a significant increase in present-moment awareness, with students who engaged in short presence exercises focused on exteroception and interoception (inner and outer sensation, respectively) reporting strong increases in their ability to sustain attention.

We also sought to account for potential baseline differences that might explain our findings. Greater mental health before the program began was associated with uptake of outward-directed social interaction and nature activities. While students who engaged more with the social interaction activity experienced negative changes in mood after program completion, they also tended to be less anxious before beginning the program, suggesting that there may have been less room for improvement in these students. This baseline effect, with less anxious students self-selecting more social interactions, could partially explain why this group ended the program feeling more depressed (e.g. through regression to the mean or range restriction). While students who focused on the quantity of social interactions started with greater mental health but felt worse after completing the program, students who felt more depressed before the program tended to engage with the inward-directed interoception micropractice. This practice, and a similar one focused on exteroception, strongly predicted increased ability to sustain attention following the program. Spirituality and resonance breathing and mindful listening stood out as areas that were not more frequently attempted by students who were feeling unwell, but showed positive associations with improved wellbeing. Resonance breathing is a form of noninvasive vagal nerve stimulation (Gerritsen and Band, 2018) that strengthens underlying physiological pathways to enhance relaxation, learning and attention (Noble and Hochman, 2019). This would presumably make it easier to automate program skills and extend them more naturally into daily life after the program ends.

Several limitations of the current study warrant consideration. First, because the data were originally collected for quality improvement purposes, this study is an exploratory post-hoc analysis. Comparisons between the *Spaces* semester and control semesters, and correlations between engagement and survey responses, do not provide evidence of a causal link between the experimental program and changes in mental health. A larger, prospective randomized trial would help strengthen our findings. It will also be important to obtain further psychometric validation of the exploratory 7-item Presence scale. An additional concern is potential ceiling effects in students who were already feeling well prior to beginning the program, which could have impacted our social interaction findings. Of note, the study population was not recruited specifically for decrements in wellbeing and/or increases in depression or anxiety, the study endpoints. However, no students reported maximal wellbeing at baseline, whereas multiple students did so on the post-program assessment. Another potential confounding variable is the reduced opportunity for social interactions as the semester progressed and students began preparing for final exams. Such a scenario could reduce wellbeing in more extroverted students while increasing it in more introverted students, representing a personality factor underlying associations between program engagement and wellbeing. Future studies should aim to better understand the impact of such potential confounders, for example by establishing student personality classifiers before program implementation. Finally, our study is limited by the self-report nature of data collection. It will be essential for future studies to combine survey data with objective measures of attention (Castelo et al., 2025) and indices of physiological function such as heart rate variability (Sevoz-Couche and Laborde, 2022; Schwerdtfeger et al., 2025). In conclusion, we argue for personalized approaches that integrate student input with built-in flexibility, since each student will engage with and responded to different elements of a wellness program.

## Acknowledgments

The authors would like to thank The Office of the University Registrar for providing class demographic data.

## Author Contributions

DJN and CLR designed and conducted the study. DJN analyzed the data and wrote the first draft of the manuscript. CLR wrote additional sections and provided critical feedback for the final version. Both authors read and approved the final manuscript.

## Data Availability Statement

These student datasets cannot be shared for legal, ethical, or privacy reasons.

## References

ACHA (2025). “American College Health Association-National College Health Assessment III: Undergraduate Student Reference Group Data Report Fall 2024”. (Silver Spring, MD: American College Health Association).

Bernardi, L., Sleight, P., Bandinelli, G., Cencetti, S., Fattorini, L., Wdowczyc-Szulc, J., et al. (2001). Effect of rosary prayer and yoga mantras on autonomic cardiovascular rhythms: comparative study. BMJ 323(7327), 1446–1449.

Bratman, G.N., Hamilton, J.P., Hahn, K.S., Daily, G.C., and Gross, J.J. (2015). Nature experience reduces rumination and subgenual prefrontal cortex activation. Proc Natl Acad Sci U S A 112(28), 8567–8572. doi: 10.1073/pnas.1510459112.

Brintz, C.E., Polser, G., Coronado, R.A., French, B., Faurot, K.R., and Gaylord, S.A. (2024). Are Formal and Informal Home Mindfulness Practice Quantities Associated With Outcomes? Results From a Pilot Study of a Four-Week Mindfulness Intervention for Chronic Pain Management. Glob Adv Integr Med Health 13, 27536130241236775. doi: 10.1177/27536130241236775.

Castelo, N., Kushlev, K., Ward, A.F., Esterman, M., and Reiner, P.B. (2025). Blocking mobile internet on smartphones improves sustained attention, mental health, and subjective well-being. PNAS Nexus 4(2), pgaf017. doi: 10.1093/pnasnexus/pgaf017.

Dajani, T., Bryant, V., Sackett, D., and Allgood, J. (2021). “Your Wellness Program Is Interfering With My Well-Being”: Reducing the Unintended Consequences of Wellness Initiatives in Undergraduate Medical Education. MedEdPublish 10.

Fincham, G.W., Kartar, A., Uthaug, M.V., Anderson, B., Hall, L., Nagai, Y., et al. (2023). High ventilation breathwork practices: An overview of their effects, mechanisms, and considerations for clinical applications. Neurosci Biobehav Rev 155, 105453. doi: 10.1016/j.neubiorev.2023.105453.

Gerritsen, R.J.S., and Band, G.P.H. (2018). Breath of Life: The Respiratory Vagal Stimulation Model of Contemplative Activity. Front Hum Neurosci 12, 397. doi: 10.3389/fnhum.2018.00397.

Hirshberg, M.J., Frye, C., Dahl, C.J., Riordan, K.M., Vack, N.J., Sachs, J., et al. (2022). A Randomized Controlled Trial of a Smartphone-Based Well-Being Training in Public School System Employees During the COVID-19 Pandemic. J Educ Psychol 114(8), 1895–1911. doi: 10.1037/edu0000739.

[Dataset] HMN (2025). Healthy Minds Study among Colleges and Universities, 2024-2025. Available: https://healthymindsnetwork.org/research/data-for-researchers.

Jain, S.J., Rakesh; Burns, Betsy (2023). WILD 5: A Proven Path to Wellness. Independently published.

Janssen, C.W., Lowry, C.A., Mehl, M.R., Allen, J.J., Kelly, K.L., Gartner, D.E., et al. (2016). Whole-Body Hyperthermia for the Treatment of Major Depressive Disorder: A Randomized Clinical Trial. JAMA Psychiatry 73(8), 789–795. doi: 10.1001/jamapsychiatry.2016.1031.

Kilrea, K.A., Taylor, S., Bilodeau, C., Wittmann, M., Gutiérrez, D.L., and Kübel, S.L. (2023). Measuring an Ongoing State of Wakefulness: The Development and Validation of the Inventory of Secular/ Spiritual Wakefulness (WAKE). Journal of Humanistic Psychology, 1– 32.

Kroenke, K., Spitzer, R.L., and Williams, J.B. (2003). The Patient Health Questionnaire-2: validity of a two-item depression screener. Med Care 41(11), 1284–1292. doi: 10.1097/01.MLR.0000093487.78664.3C.

Kuyken, W., Ball, S., Crane, C., Ganguli, P., Jones, B., Montero-Marin, J., et al. (2022). Effectiveness and cost-effectiveness of universal school-based mindfulness training compared with normal school provision in reducing risk of mental health problems and promoting well-being in adolescence: the MYRIAD cluster randomised controlled trial. Evid Based Ment Health 25(3), 99–109. doi: 10.1136/ebmental-2021-300396.

Noble, D.J., Goolsby, W.N., Garraway, S.M., Martin, K.K., and Hochman, S. (2017). Slow Breathing Can Be Operantly Conditioned in the Rat and May Reduce Sensitivity to Experimental Stressors. Frontiers in Physiology 8. doi: 10.3389/fphys.2017.00854.

Noble, D.J., and Hochman, S. (2019). Hypothesis: Pulmonary Afferent Activity Patterns During Slow, Deep Breathing Contribute to the Neural Induction of Physiological Relaxation. Front Physiol 10, 1176. doi: 10.3389/fphys.2019.01176.

Rolin, D., Fox, I., Jain, R., Cole, S.P., Tran, C., and Jain, S. (2020). Wellness Interventions in Psychiatrically Ill Patients: Impact of WILD 5 Wellness, a Five-Domain Mental Health Wellness Intervention on Depression, Anxiety, and Wellness. J Am Psychiatr Nurses Assoc 26(5), 493–502. doi: 10.1177/1078390319886883.

Schwerdtfeger, A.R., Tatschl, J.M., and Rominger, C. (2025). Effectiveness of 2 Just-in-Time Adaptive Interventions for Reducing Stress and Stabilizing Cardiac Autonomic Function: Microrandomized Trials. J Med Internet Res 27, e69582. doi: 10.2196/69582.

Sevoz-Couche, C., and Laborde, S. (2022). Heart rate variability and slow-paced breathing:when coherence meets resonance. Neurosci Biobehav Rev 135, 104576. doi: 10.1016/j.neubiorev.2022.104576.

Spitzer, R.L., Kroenke, K., Williams, J.B., and Lowe, B. (2006). A brief measure for assessing generalized anxiety disorder: the GAD-7. Arch Intern Med 166(10), 1092–1097. doi: 10.1001/archinte.166.10.1092.

Stuckey, H.L., and Nobel, J. (2010). The connection between art, healing, and public health: a review of current literature. Am J Public Health 100(2), 254–263. doi: 10.2105/AJPH.2008.156497.

Topp, C.W., Ostergaard, S.D., Sondergaard, S., and Bech, P. (2015). The WHO-5 Well-Being Index: a systematic review of the literature. Psychother Psychosom 84(3), 167–176. doi: 10.1159/000376585.

VanderWeele, T.J., Johnson, B.R., and Bialowolski, P.T. (2025). The Global Flourishing Study: Study Profile and Initial Results on Flourishing. Nat. Mental Health. doi: 10.1038/s44220-025-00423-5.

Waldinger, R.J., and Schulz, M.S. (2023). The good life : lessons from the world’s longest scientific study of happiness. New York: Simon & Schuster.

