## Supplementary Material for "*Spaces*: A Student-Informed, Course-Based Program to Enhance Wellbeing and Present-Moment Awareness in University Undergraduates"

### 1 Table S1. Class Demographics and Scale Averages

| | **CONTROL SEMESTERS** F20, F21, S22[b], S24, S25[c] | ***SPACES*** **SEMESTER** F24 | **AVG/TOT**[a] (% of full data set) |
|---|---|---|---|
| **Class Composition** | | | |
| Freshman (% of class[es]) | 21 (7.8) | 0 (0.0) | 21 (7.0) |
| Sophomore (% of class[es]) | 67 (24.8) | 3 (10.0) | 70 (23.3) |
| Junior (% of class[es]) | 82 (30.4) | 11 (36.7) | 93 (31.0) |
| Senior (% of class[es]) | 100 (37.0) | 16 (53.3) | 116 (38.7) |
| Total Enrolled (% of full data set) | 270 (90.0) | 30 (10.0) | 300 |
| **Ethnicity and Legal Sex**[d] | | | |
| Asian | 36 (28.3) | 5 (16.7) | 41 (26.1) |
| Black | 30 (23.6) | 7 (23.3) | 37 (23.6) |
| Hispanic | 18 (14.2) | 5 (16.7) | 23 (14.6) |
| Multi | 1 (0.8) | 1 (3.3) | 2 (1.3) |
| Unknown | 1 (0.8) | 2 (6.7) | 3 (1.9) |
| White | 41 (32.3) | 10 (33.3) | 51 (32.5) |
| Female | 93 (73.2) | 25 (83.3) | 118 (75.2) |
| **Average Scores on Wellness Scales** | | | |
| GAD-7 (mean ± SD) | 7.53 ± 0.82 | 5.82 ± 0.64 | 6.67 |
| PHQ-2 (mean ± SD) | 1.73 ± 0.25 | 1.14 ± 0.17 | 1.43 |
| WHO5 (mean ± SD) | 11.58 ± 0.79 | 15.06 ± 0.90 | 13.32 |

***[a]AVG/TOT = average and total***
***[b]2 Seniors included in total withdrew***
***[c]1 Junior and 1 Senior included in total withdrew***
***[d]Ethnicity and Legal Sex for control semesters is broadly representative; does not include all students due to incomplete university datasets***

### 2 Result S1. Exploratory Analysis of Engagement Profiles Corresponding to Mental Health Outcomes

A secondary exploratory analysis sought to identify activities and practices from the *Spaces* program that predicted improvements in anxiety, depression, and wellbeing. For this analysis, area under the curve (AUC) was calculated for each wellness scale for each student from the starting point of the program to the end of the semester, with individual baseline values standardized to zero. Students were divided roughly into thirds (categories of high, medium, and low AUC) with high AUC corresponding to the greatest improvements in mental health over the program period (thus negative for the GAD-7 and PHQ-2 and positive for the WHO5). Times spent on each practice were compared between students with high vs. low AUC for an exploratory classification of engagement predictors. Ratios were determined by dividing engagement values (self-reported and converted to minutes) in the high by the low AUC groups.

**Figure S1** below plots relative engagement ratios for each scale (i.e. approximate minutes students engaged with each program activity or practice in the high versus low improvement groups). Noteworthy descriptive results, categorized by scale, were:

<u>GAD-7:</u> Students in the high AUC group (i.e. with reduced anxiety from baseline over the program period) devoted 2.02x as much time to *Mindful Listening* and 1.87x as much time to *Spirituality and Resonance Breathing*. They tended to be less likely to engage with *Mindful Action* (0.40x), *Intentional Interactions* (0.46x) and *Nature* (0.53x).

PHQ-2: Students in the high AUC group (i.e. with reduced depression from baseline over the program period) devoted 2.43x as much time to *Exteroception*, 1.64x as much time to *Interoception*, and 1.51x as much time to *Nature*. They tended to be less likely to engage with *Intentional Interactions* (0.65x), *Creative Hobbies* (0.68x), and *Social Media Detox* (0.69x).

WHO5: Students in the high AUC group (i.e. with increased wellbeing from baseline over the program period) devoted more time to *Mindful Listening* (1.84x) and *Adaptive Stress* (1.51x), although there was a wide range of overrepresented practices (**Figure S1**, *right*). *Creative Hobbies* (0.55x) *Nature* (0.55x), and *Intentional Interactions* (0.59x) were underrepresented.

Summary/Interpretation: Unique clusters of engagement with program components differentially associated with cumulative improvements in anxiety, depression, and overall wellbeing over the *Spaces* program weeks. While our preliminary analysis lacked statistical power to detect differences due to large interindividual variability, these engagement clusters suggest that over the program period: 1) individuals who engaged more with spirituality and resonance breathing, and mindful listening, tended to feel less anxious, 2) individuals who engaged more with short and simple mindfulness practices focused on interoception and exteroception in nature tended to feel less depressed, 3) an array of activities and practices including mindful listening, adaptive stress, spirituality and resonance breathing, and social media detox tended to associate with improved overall wellbeing, and 4) only intentional interactions predicted decreased student wellness across all scales.

### 3 Figure S1. Exploratory Engagement Profiles for Reduced Anxiety and Depression, and Increased Wellbeing

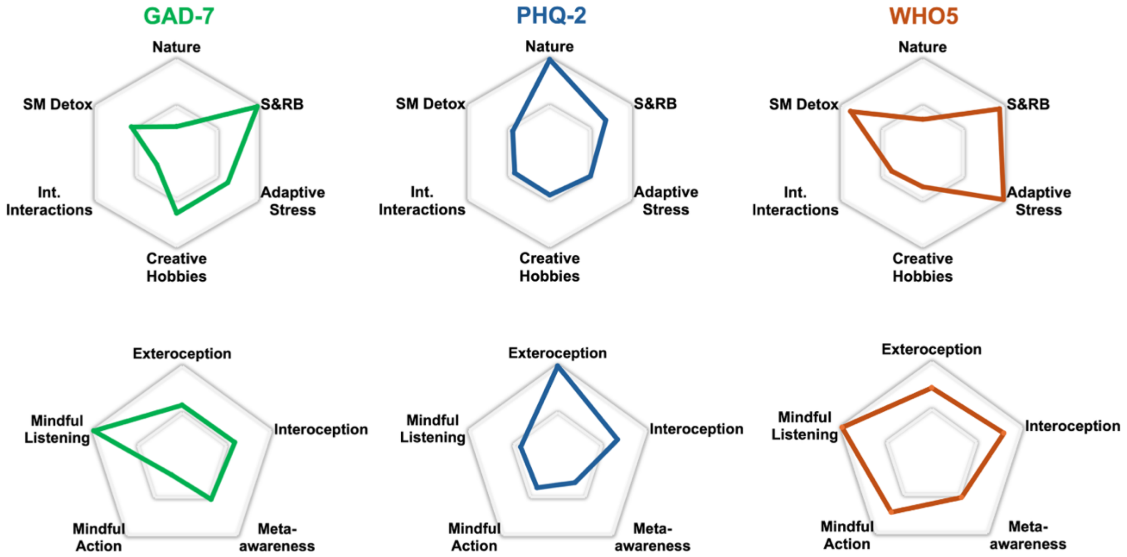


Area under the curve (AUC) was calculated for each wellness scale over the program timeline, with higher values indicating students who became less anxious (GAD-7) or depressed (PHQ-2), or more well (WHO5) over the program period. Engagement with specific program activities (*top*) and presence micropractices (*bottom*) categorized students who saw the biggest improvements in different facets of mental health. The corners of each polygon represent engagement ratios (high/low AUC) for individual program components, i.e. the extent to which students who felt better after the program tended to engage with that activity or practice compared to those who felt worse. The outer perimeter of each subpanel is standardized to 1 for visual clarity. See **Result S1** for further details and **Table 1** for a description of program components. *Int. Interactions, Intentional Interactions; S&RB, Spirituality & Resonance Breathing; SM Detox, Social Media Detox*

### 4 Table S2. Survey Instrument[a]

| |
|---|
| **GAD-7:** "Over the **last week**, how often have you been bothered by the following problems?"<br><br>*0 — Not at all; 1 — Several days;*<br>*2 — More than half the days; 3 — Nearly every day*<br><br>**Q1. Feeling nervous, anxious, or on edge**<br>**Q2. Not being able to stop or control worrying**<br>**Q3. Worrying too much about different things**<br>**Q4. Trouble relaxing**<br>**Q5. Being so restless that it's hard to sit still**<br>**Q6. Becoming easily annoyed or irritable**<br>**Q7. Feeling afraid as if something awful might happen**<br><br>*Recommended Cutoff:* **10 or greater** |
| **PHQ-2**: "Over the **last week**, how often have you been bothered by the following problems?"<br><br>*0 — Not at all; 1 — Several days;*<br>*2 — More than half the days; 3 — Nearly every day*<br><br>**Q1. Little interest or pleasure in doing things**<br>**Q2. Feeling down, depressed, or hopeless**<br><br>*Recommended Cutoff:* **3 or greater** |
| **WHO5:** "Please indicate how you have been feeling over the **last week** for each of the five statements."<br><br>*0 — At no time; 1 — Some of the time; 2 — Less than half of the time;*<br>*3 — More than half of the time; 4 — Most of the time; 5 — All of the time*<br><br>**Q1. I have felt cheerful and in good spirits**<br>**Q2. I have felt calm and relaxed**<br>**Q3. I have felt active and vigorous**<br>**Q4. I woke up feeling fresh and rested**<br>**Q5. My daily life has been filled with things that interest me**<br><br>*Recommended Cutoff:* **below 13** |

**Presence (Wks. 8 & 13):** "To what extent have the following statements been characteristic of or true for you, **over the last week**?"

*1 — Strongly disagree; 2 — Disagree;*
*3 — Neutral/unsure; 4 — Agree; 5 — Strongly agree*

**Q1. I have felt a background sense of peace**
**Q2. Life has flowed with harmony and ease**
**Q3. My senses and the world around me have seemed vivid and alive**
**Q4. I have enjoyed performing daily activities and having ordinary experiences**
**Q5. I have noticed arising thoughts and emotions without following them 'down the rabbit hole'**
**Q6. My interactions with others have gone smoothly and I have refrained from instant reactions or judgements**
**Q7. I have been able to focus my attention on something for long periods of undistracted time**

**Engagement (Wk. 13):** "Please indicate **approximately how many minutes per day** you spent trying out the program activities or practices listed below over the last 30 days, specifically because of this program/assignment (i.e., for this question, do not count activities or practices that were already part of your life before beginning the program unless you intentionally devoted more time to them). *For social media detox, indicate the approximate time* ***removed*** *from social media or technology use per day."*

*0 minutes; 1-5 minutes; 6-10 minutes;*
*11-20 minutes; 21-30 minutes; More than 30 minutes*

**Q1.** Nature activity
**Q2.** Spirituality & Resonance Breathing activity
**Q3.** Adaptive Stress activity
**Q4.** Creative Hobbies activity
**Q5.** Quality Relationships activity - intentional interactions
**Q6.** Quality Relationships activity - social media detox
**Q7.** Exteroception practice
**Q8.** Interoception practice
**Q9.** Meta-awareness practice
**Q10.** Mindful Action practice
**Q11.** Mindful Listening practice

**Q12. Average time spent trying out all program activities or practices each day:**
*0 minutes; 1-10 minutes; 11-20 minutes; 21-30 minutes;*
*31-45 minutes; 46-60 minutes; More than 60 minutes*

**Q13. I accomplished these averages by engaging with the program:** *fairly consistently throughout the 30 days; mostly during the first 15 days; mostly during the last 15 days; barely or not at all; other (please specify in survey comments)*

[a]***Scales were administered in the Canvas Learning Management System; students used dropdown menus to select answers for each question.***
